# Model calibration, nowcasting, and operational prediction of the COVID-19 pandemic

**DOI:** 10.1101/2020.04.14.20065227

**Authors:** J. D. Annan, J. C. Hargreaves

**Affiliations:** Blue Skies Research Ltd, The Old Chapel, Albert Hill, Settle, BD24 9HE, UK

## Abstract

The COVID-19 pandemic has already caused huge human and economic cost, and will likely continue to do so for some time. Accurate and timely forecasting is essential in order for policy-makers to anticipate future requirements so that they may plan and respond effectively. We present a model-based nowcasting and forecasting system based on calibration of a dynamical epidemic model to observational data. We apply Bayesian model calibration and initialisation methods that are well established in other fields, but that do not appear to have been used in the UK in the current outbreak. We show that it would have been possible using our methods to diagnose and forecast the rapid growth of the COVID-19 outbreak in the UK by mid-March. We also show that model calibration can successfully estimate the effect of the lockdown policies in various European countries, deducing correctly that the reproduction number was likely less than 1 around three weeks after the lockdown date. This is significantly earlier than has been achieved by other approaches. Our current (16 May) estimate for the reproduction number in the UK is *R* = 0.8 with a 95% range of 0.7−0.9.

## Introduction

The COVID-19 pandemic has caused huge disruption, with enormous human cost and economic damage. By the 10th of May 2020, over 250,000 lives have been lost globally and while it is surely too early to assess the final outcome, the financial cost of the pandemic so far has been estimated at several percent of global GDP amounting to trillions of dollars [1]. In the UK, over 30,000 fatalities have been recorded.

In order for model forecasts to be relevant and useful, they must be calibrated correctly so as to adequately represent the situation they are attempting to simulate. The term calibration as used here refers generally to the process by which model inputs (parameters and initial state variables) are tuned via an automatic algorithmic process in order that the model outputs provide a reasonable match to observational data relating to a specific application, such as simulating an outbreak in a particular region [2,3]. The successes of such model-based forecasting in epidemiology has been well documented [4–7], but the process of calibration still seems rare in practice [8]. While some forecasts based on calibrated dynamical models have been produced for the recent COVID-19 outbreak in some regions [9], we are unaware of any that focussed on the UK during the early stages of the epidemic.

We model the outbreak using a simple SEIR model and use a Bayesian approach to calibrate the main parameters of the model to observational data. The method is outlined in the following section. We demonstrate the importance and performance of our approach through two main applications in this paper. We firstly consider the initial forecasting of the epidemic in its early stages. We show that our model can be effectively calibrated at an early stage of the outbreak so as to accurately predict its future growth, reproducing the very rapid exponential growth that was seen in the UK throughout March. We contrast this with the performance of a more complex but uncalibrated model which was used by the Scientific Pandemic Influenza Group on Modelling (SPI-M) to forecast the outbreak and which provided guidance to the Scientific Advisory Group for Emergencies (SAGE). Secondly, we consider the use of model calibration for assessment of the effectiveness of recent policy interventions aimed at suppressing the epidemic, and ongoing short-term forecasting of the epidemic’s progress. We validate our approach through hindcasting the outbreak in Hubei Province, China before going on to apply our approach to the UK and several other European countries, showing that the calibrated dynamical model appears to outperform several statistical analyses in in estimating the current reproductive number and in forecasting over the near term. We deduce that suppression has been achieved in all of these countries, and demonstrate that this was detectable much earlier using our method than it was with the alternatives analyses.

## Materials and methods

### Model

We use a simple unstratified SEIR model for our simulations. While this model appears surprisingly capable for the purposes of generating useful analyses and forecasts, we expect that more sophisticated and detailed models that incorporate age and social structure (but which were not available to us) would be capable of even better results if appropriately calibrated. In our model, the total population (considered as a continuum) is divided into Susceptible, Exposed, Infectious, and Recovered groups respectively, and moves between these groups according to the standard equations:

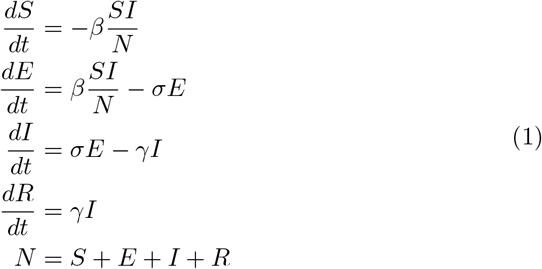

where the rate parameters *β = R*_0_*/τ_I_*, *σ =* 1*/τ_L_* and γ = 1**/***τ_I_* are defined in terms of the reproductive number *R*_0_ and latent and infectious periods *τ_L_* and *τ_I_* respectively. Performing a model simulation requires that these parameter values, and the initial state (e.g. infected proportion proportion), are defined at the initial time *t* = 0. We model death with an uncertain infection fatality rate (IFR), and a time-to-death function applied to the infectious population [10].

### Uncertain parameters

We consider a total of 6 uncertain parameters, which are listed in Table 1. For calibration of the model to the early stage of the epidemic, all model parameters are considered constant in time. When we consider longer time frame which includes interventions which were being introduced to reduce transmission, the reproductive number is considered piecewise constant, changing instantaneously from its initial value, *R*_0_, to some new value, *R_t_*, at the time of the intervention. This piecewise constant approach is similar to that of other research [10], but for simplicity we only consider a single intervention date for each country to represent the net impact of all interventions. For the initial reproductive rate we use a Gaussian prior of N(3,1^2^) to represent a wide range of plausible values [9, 11,12], and for *R_t_* we use N(1,0.5^2^) in in order to represent the expectation that the controls will have a substantial effect albeit we are uncertain whether these achieve suppression (*R_t_* < 1) or merely mitigation (*R*_0_ *> R_t_ >* 1). Using a prior centred on unity means that one can easily discern from the posterior distribution whether the observed data are more consistent with suppression or mitigation even before the effect of the policies is known with high confidence.

**Table 1.**
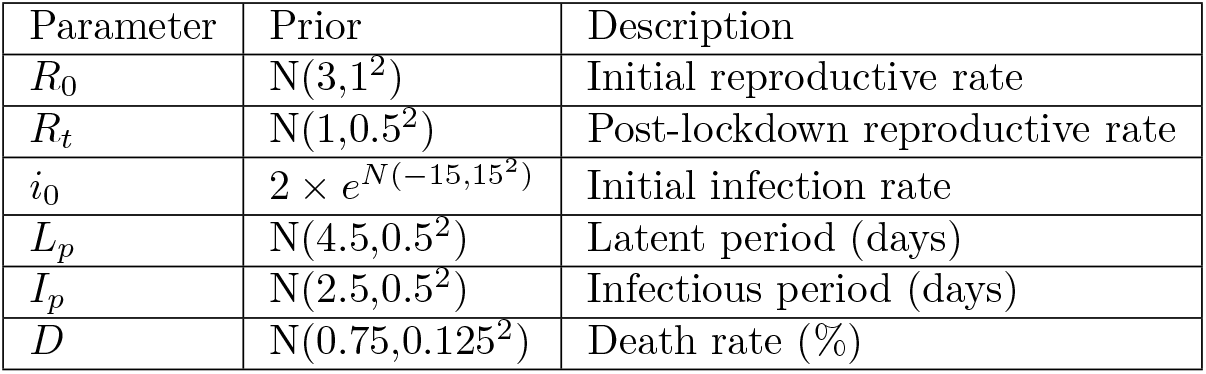
Tunable parameters and priors.

The starting size of the epidemic is taken to be uncertain with a very broad prior that ensures the specification of start date is not critical so long as a reasonable choice is made. For the latent and infectious periods we use priors of N(4.5,0.5^2^) and N(2.5,0.5^2^) respectively, giving a prior mean serial interval of 5.75 days and a 95% range of 4.7–6.8 days [11–13]. The infection fatality rate was set at N(0.75%,0.125%^2^), consistent with estimates for a case fatality rate of around 1.5% [14, 15] and an assumption that roughly half of all infections are broadly asymptomatic and thus undiscovered [16]. Our judgement in this was subsequently supported by a meta-analysis [17] which was published as we were completing this work. The start date of each simulation was fixed at a few weeks before cases were first observed: the results are insensitive to this choice due to the broad prior on the size of initial infection.

### Data

For modelling European countries (including the UK up to late April), we used the median of three sources [18–20], as we found no single source to be completely reliable in their reporting of the official statistics. Occasional missing days were filled by smoothing. For the later analyses of UK data after the switch in reporting standards discussed later, we rely solely on the official UK data (https://coronavirus.data.gov.uk/). For the analyses of interventions, we also use regional data for Hubei, China which were obtained from https://www.kaggle.eom/imdevskp/corona-virus-report#covid_19_clean_complete.csv. This data set did not include the earliest deaths and these were added by hand based on the Wikipedia page: https://en.wikipedia.org/wiki/2019-20_coronavirus_pandemic_in_mainland_China. Some further limitations of the data are discussed where they are used.

### Calibration method

Estimation of the initial state of the model, and the choice of its parameter values, are sometimes considered as two distinct steps in forecasting using computer models, but while there are often practical reasons for this there is no fundamental mathematical distinction between these two tasks. In this work we jointly estimate them in a single calibration process and use the term “state” in a broad sense to include model parameter values as well as prognostic variables. Rather than providing a detailed and complex methodological description our intention here is merely to provide a very brief introduction and summary to a general audience, and then show compelling motivation for the broader use of these techniques, through demonstrating the large benefits which it could have brought to our understanding and management of the current epidemic of COVID-19.

Calibration is performed in a Bayesian probabilistic sense. In the Bayesian paradigm, we use the language and mathematics of probability to describe our uncertain knowledge of the world. Bayes’ Theorem describes how a prior estimate *p*(Θ) of the state Θ can be updated in the light of observational evidence *O* according to the equation:

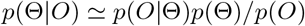

In the above, *p*(Θ|*O*) is the posterior estimate of the state conditioned on the evidence, *p*(Θ) is our prior belief before accounting for the evidence, and *p*(*O*|Θ) is the likelihood function that calculates the probability of obtaining the particular observations *O*, as a function of the state variables/parameters Θ. The likelihood calculation is where the model is confronted with reality and is outlined in the following section. Calibration was performed using a Markov Chain Monte Carlo (MCMC) algorithm [21, 22] using the ‘MCMCPack’ package in the R language. The MCMC algorithm performs a random walk in parameter space in order to sample the posterior, at each step generating the model prediction resulting from that set of parameters, and evaluating the likelihood function (described below) in order to determine the probability of obtaining the observations, given these parameter choices. The algorithm outputs the posterior probability distribution in the form of an ensemble of equally probable parameter sets which sample this distribution. Code to generate all results and figures presented here is included as supplementary information.

### Likelihood function, observational uncertainties and model error

In order to calculate the likelihood *p*(*O*|Θ), we must account for the uncertainties in how the observations are generated from the underlying reality. We consider three major sources of uncertainty. The model-data comparison is performed in log-space due to the largely log-linear dynamics, and all of the following terms are treated as multiplicative errors on the model output (additive errors in log space). To handle the obvious problem with observations of zero close to the start of the epidemic, we replace these values (which are rare) by values of 0.5.

Firstly and most straightforwardly, sampling uncertainty arises due to the stochastic nature of deaths occurring in a specific interval such as a day. Since we are using lightly processed data with some non-integer values we use a Gaussian approximation 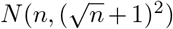 to the Poisson distribution for n expected cases or deaths, with the additional +1 included to ensure adequate uncertainty at very low values of *n* and to minimise the influence of additional cases and deaths which were added where zero were observed.

A second source of uncertainty is reporting errors. These have been much discussed in the UK context and there is clear evidence of them having a weekly pattern due possibly to working practices in the medical sector. In particular, the “weekend dip” throughout April has been substantially greater than could plausibly be accounted for via sampling uncertainties alone and therefore as our second source of uncertainty we include an additional factor for these reporting errors, for which we use a Gaussian distribution with a magnitude of 20% of the expected value (again at one standard deviation).

Finally, perhaps the most subtle and important source of uncertainty is the inadequacy of the model itself. Even if the model is perfectly initialised, with the best possible choice of parameter values, it is inevitable that it will diverge from reality to a greater extent than could be accounted for by stochastic factors. There are in fact no ‘true’ constant parameter values for which the model can track reality indefinitely, as the instantaneously effective values of the rate parameters in Equation 1 will vary through time. In particular *β*, which depends on the reproductive number, will change as the disease spreads among different sections of the population, and it may also be affected by external factors such as climatic conditions and behavioural changes beyond the scope of our model. We may also expect the average IFR to vary according to which sectors of the population are most heavily infected at any one time. This failure of the model to perfectly represent reality does not just affect the forecast, but also prevents the model from ‘shadowing’ [23] an indefinitely long historic time series of observations. Thus, model inadequacy must be considered as intrinsic to the calibration process [24]. Here we treat the model inadequacy in a straightforward manner as a multiplicative error of 5% per day, equivalent to an irreducible uncertainty in *R*_0_ (and *R_t_* where used) of this magnitude. This model inadequacy term is necessarily a rather subjective matter and experts might be able to implement a more sophisticated approach to this, though it is inevitably hard to quantify. We can take some guidance from an assessment of epidemic forecasts [7], which showed significant skill from similar modelling (albeit in a very different scenario) over a fortnightly time scale. An error term of 1% per day or less would not allow for significant model error within the time scale of interest, whereas to use a value of 10% per day or greater would be to assert that the model was rather poor with a limited forecast horizon. This model error grows both forwards and backwards in time from the ‘present’, i.e. the forecast date. What this means in practice is that historical observations sufficiently distant in time from the present have little influence on the calibration, and that the posterior uncertainty on parameter values will remain bounded below even for an unlimited time series of observations.

These three error terms are independent, Gaussian and additive in log-space and therefore are added in quadrature to give a total uncertainty on model predictions which then allows us to calculate the probability of obtaining the observations from a given model simulation. We emphasise that the approach taken here is only intended as a simple benchmark that experienced epidemiological modellers will hopefully be able to surpass.

## Case study 1: Initial epidemic growth

In our first case study, we consider the issue of forecasting the initial growth of the outbreak in the UK. In the early stage of community spread, an epidemic exhibits exponential growth at a constant rate 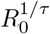 which depends on the reproduction number *R*_0_ and the reproductive time scale of the disease which we denote as *τ*. On 16th March, by which time community spread was established, forecasts of the future progress of the pandemic in the UK under a range of scenarios were published [13]. The model parameters in these simulations included an *R*_0_ parameter of 2.4 (with a range of 2.0–2.6 tested in sensitivity analyses) and a serial interval of *τ* = 6.5 days. Together these parameters imply a doubling time of just over 5 days, with the sensitivity tests representing doubling times ranging from 4.7 to 6.5 days. These values are consistent with the assessment of SPI-M-O/SAGE up to the 18th March that *R*_0_ was in the range of 2.0–2.5 [25] and that the doubling time was around 5–7 days [26]. While the model used in that work represents the state of the art and contains multiple compartments representing different age categories and social groups, its basic behaviour at national scale — as portrayed in their manuscript — can be well simulated by our simpler SEIR model as we show below in Figure 2B.

Using our Bayesian approach, we calibrated the SEIR model to the daily case numbers for the UK between March 1st (this being the first day where 10 cases were observed) to the 14th, in order to focus on the interval in which sustained community transmission was taking place. Testing policy in the UK was still consistent and arguably adequate to represent the growth in infections, with the Government pursuing a containment policy up to the 12th March, so we would expect these figures to provide a useful representation of the true growth rate of the epidemic even with reported cases being a likely undercount of the total infection numbers. When the time series covers several doubling times (as is the case here), moderate changes in the case ascertainment rate will not lead to an unacceptably large error in apparent growth rate, and it is more likely that growth rates will be underestimated than overestimated [27]. The mean posterior estimate for *R*_0_ we obtained from calibrating to this data set is 3.5 with a 95% range of 2.3–4.7 as shown in Figure 1A. When taken in conjunction with our posterior estimates of latent and infectious periods, the resulting doubling time is found to be 2.8 days with a 95% range of 2.3–3.7 days, roughly half that of the model results presented to SAGE. The growth rate depends not only on *R*_0_ but also on the serial interval [28], and these two factors are strongly correlated in our posterior. The doubling time is therefore much more tightly constrained than the *R*_0_ parameter itself. In order to facilitate a clearer comparison with the parameters used by the modelling presented to SAGE we repeated the calibration process but imposed the mean serial interval of 6.5 days (by using fixed values of *L_p_* = 5 days and *I_p_* = 3 days in our model). In this case, our mean posterior estimate for *R*_0_ increases to 4.0 with a 95% range of 2.9–5.1, giving essentially the same estimated doubling time as before of 2.9 days (95% range 2.4–3.7 days). Thus when considered either in terms of doubling time, or *R*_0_ value using the same serial interval, our calibrated posterior is very different to the values modelled for SAGE.

**Fig 1.**
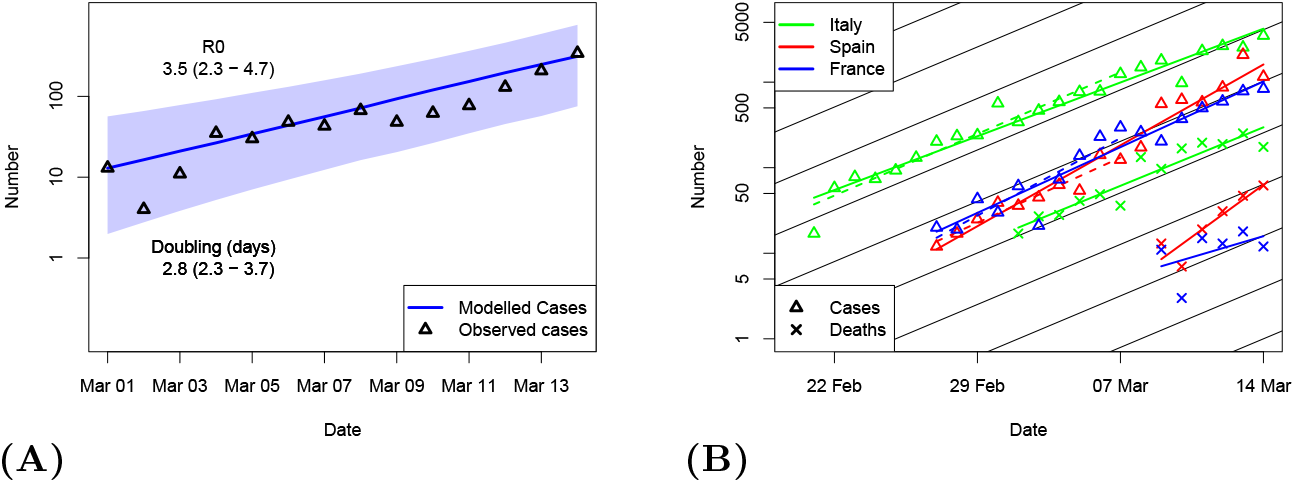
Calibration of the model. A Calibration of model to reported case number in UK. Blue line and plume is median and 95% range of posterior. Black triangles indicate data points. B Cases (triangles) and deaths (crosses) from three European countries as indicated by colour. Solid coloured lines are least squares fits to all data up to 14 March. Dashed coloured lines are least squares fits to case data up to 7 March. Black lines show slope of 3 day doubling.

Data from Italy, Spain and France (which were all some way ahead of the UK in terms of development of their outbreaks) also pointed strongly towards doubling time in the region of 3 days by mid-March. Specifically, simple linear least squares fits to the logarithm of the daily case number data in Italy, Spain and France give doubling times of 3.3, 2.1 and 2.7 days respectively, and equivalent analyses on their more limited daily death data give doubling times ranging from 1.7 to 4.3 days (Figure 1B, solid coloured lines). The results from the case number data are barely changed if we only use data up to 7th March (Figure 1B, dashed coloured lines). More comprehensive analyses of European data [27, 29] also diagnose doubling times of around 3 days with the latter describing the evidence for this as “overwhelming.” Although both these papers used data a little way past the cut-off date of 14th March considered here, it is hard to avoid the conclusion that the evidence was at least strong by that time.

Model projections made with the calibrated (posterior mean) parameters and uncalibrated models are presented for validation in Figure 2A. Beyond mid-March, testing policy changed abruptly in the UK and case numbers fell rapidly behind the true infection number, so we use death data for validation, and only consider observations up to early April, prior to any influence from the major policy interventions relating to social distancing which were imposed in the UK by the 23rd March. For clarity in these comparisons, we only display the posterior mean from our calibration. While the initial growth rate in the noisy and limited death data up to 14th March appears consistent with our calibrated model, we do not perform any further calibration with this time series and merely use the total number of deaths up to the 14th March to time the start of both simulations [13]. Deaths up to that date are therefore identified as initialisation data in the figure. Our calibrated simulation generates a prediction that matches the subsequently observed data remarkably well. In contrast, the uncalibrated model with a 5-day doubling time performs poorly, underestimating the true daily death number by a factor of more than 10 at the end of March, a mere two weeks after the simulation was published. Figure 2B shows the full forecasts for the epidemic in the case of no intervention. The accuracy of our replication of the more complex model [13] can be seen from their predicted death curve (their Figure 2) which is very similar to our uncalibrated results, having a peak in late May perhaps one week earlier than our replication, and with our maximum of just under 16k deaths per day comparing closely to their value of 15k. The superior performance of the simple model, when properly calibrated, over a state of the art model, shows the importance of calibration.

**Fig 2.**
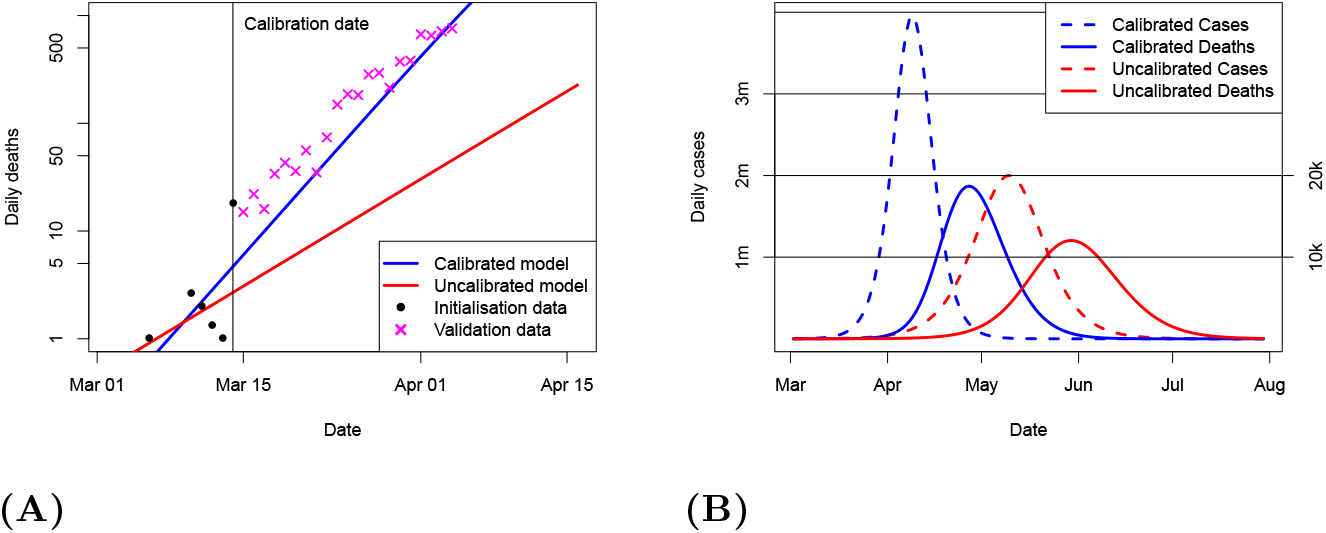
Calibration and forecasts of the UK outbreak. A Short-term forecast validation. Lines show calibrated and uncalibrated model deaths. Data points indicated by black dots (initialisation) and magenta crosses (validation). B Full unmitigated epidemic forecasts of calibrated (blue) and uncalibrated (red) models, for both daily infections (dashed, left hand scale) and deaths (solid, right hand scale).

The calibrated forecast predicts a peak of over 3 million new cases per day, nearly twice the height of the uncalibrated case. Perhaps more critically, the timing of this peak is brought forward by a full month, to early in April, which was less than a month after the date of issue of the forecast. Official advice given by SAGE as late as 18 March predicted a 5–7 day doubling time [26] and the lockdown policy was not implemented until after this was corrected several days later [30]. It is clearly not necessary to perform a formal model calibration in order to discover the rapid doubling time exhibited by the data, but it is only through the use of calibrated models that useful forecasts can be made to inform and enable planning for the impact of the epidemic and the policy response to it. We also note that a delay of 1 week in suppressing the epidemic, in the context of a doubling time of around 3 days, will lead to an epidemic peak (and total number of cases and deaths) that is around 5 times (ie 2^7/3^) larger than would have been the case with more prompt action. Equivalently one could say that acting one week sooner could have reduced the size of the outbreak by around 80%, with an uncertainty range of 73%–88% based on the estimated range of doubling times arising from our model calibration.

## Case Study 2: Evaluation of the lockdown, and short-range forecasting

For our second case study, we consider analysis of the effectiveness of policy interventions and the potential for short-term forecasts following such interventions. Policies have been put in place in the UK and other countries to limit social contact with the aim of suppressing or at least mitigating the outbreak. We model this via a reduction in the reproductive number from its initial value *R*_0_ to some lower value which we denote as *R_t_*. We use the term suppression to refer to the case where *R_t_* < 1 and the outbreak immediately declines, versus mitigation where *R_t_* is reduced from its original value *R*_0_ but remains in excess of unity, meaning that the outbreak will still grow (until we reach a state of herd immunity relative to this *R* value) but more gradually and to a lower total extent. Our aims here are twofold: we estimate the values of *R*_0_ and *R_t_* (jointly with other parameters and initial state as before), and we also use these parameter estimates to forecast the future trajectory of the outbreak with our model. A similar approach has been presented using a statistical model [10].

It is clear that during the large scale uncontained outbreaks observed in multiple countries the reported case numbers become increasingly unreliable, with the most plausible estimates of infection number appearing to arise from inverse calculations which use the observed deaths and an assumed mortality rate [14] and therefore we do not use the case number data for calibration, relying solely on daily reports of death numbers. We use daily reports of death numbers in a range of European countries to simultaneously estimation 5 model parameters and the initial state. In the next section we validate this approach using data from Hubei, China.

## Validation in Hubei

While our main interest is in the UK, the initial outbreak in the city of Wuhan in Hubei province, China, provides the most comprehensive test of the long-term forecasting performance throughout the epidemic due to the long time series available. Figure 3 shows the results of calibration to various initial subsets of the data together with the resulting forecasts, performed at three-weekly intervals starting 10 days after the lockdown date.

**Fig 3.**
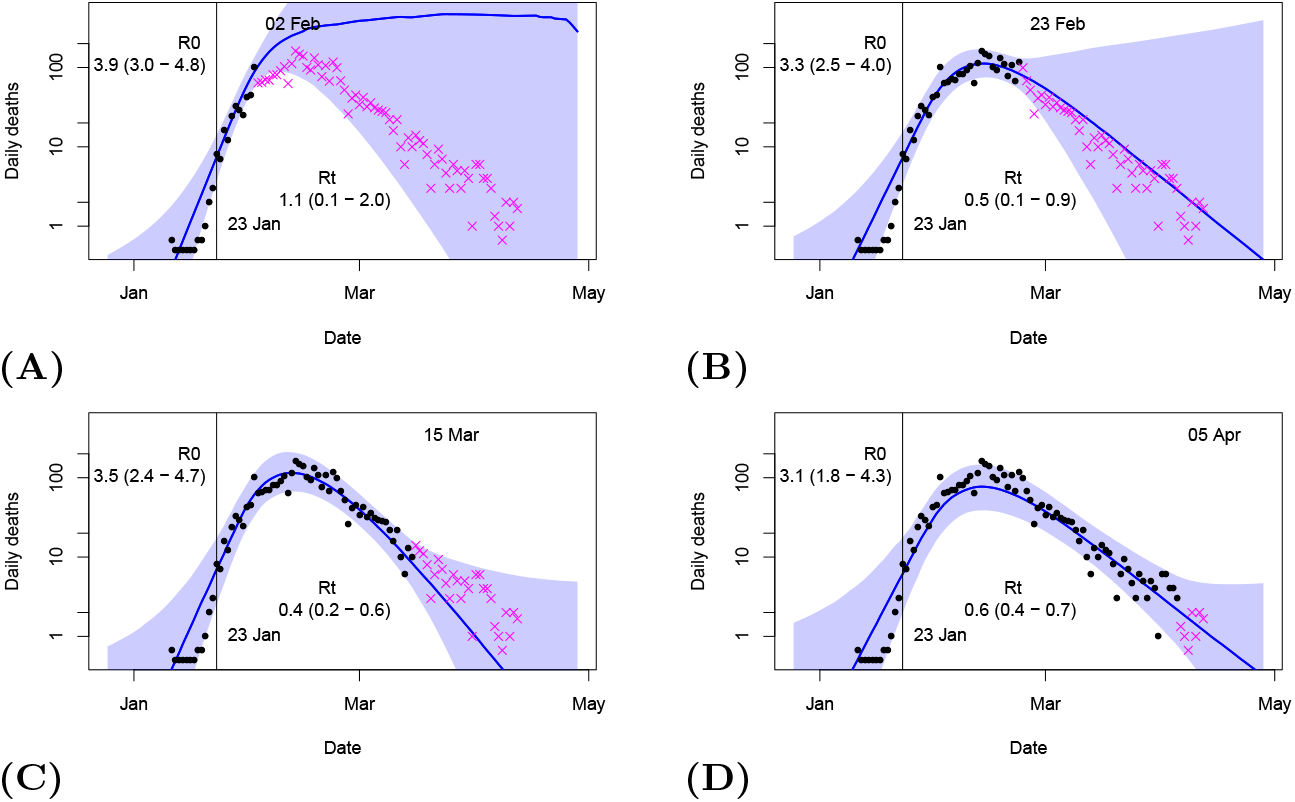
Hindcasts and forecasts from Hubei, China. Forecasts issued at three-week intervals on dates shown. Median hindcast/forecast is shown as blue line, with blue plume as 95% credible interval. Date of lockdown shown as vertical line, with estimated *R*_0_ and *R_t_* values shown on plots as mean and 95% ranges. Black dots are daily deaths which were used for calibration of each forecast. Magenta crosses are validation data observed subsequently to forecast date.

Figure 4 shows the estimates of *R_t_* corresponding to weekly forecasts, including the four cases shown in Figure 3. For the first two forecasts on Figure 4, at 3 and 10 days post-lockdown, the observational data on death numbers provides no constraint on *R_t_*, as there has not been sufficient time for any changes in infection rate to affect the number of deaths. Therefore, the estimate of *R_t_* is simply the prior range and the corresponding forecast in Figure 3A reflects this high level of uncertainty. By 17 days, however, there is clear evidence — albeit not overwhelming — for *R_t_ <* 1, and confidence in the success of suppression exceeds 97.5% by the time of the 31 day analysis and stays above that level thereafter. The forecasts are reasonably well nested, as would be expected in a well designed Bayesian system that learns from new information, and the final posterior median is contained in all prior estimates. With a total of 8 data points lying outside the forecast plumes and 151 (including repetitions) lying inside, it appears that our forecasts are reasonably well calibrated in the technical sense of forecast uncertainty matching observed spread reasonably well. While we should not read too much into one example, this level of performance seems encouraging.

**Fig 4.**
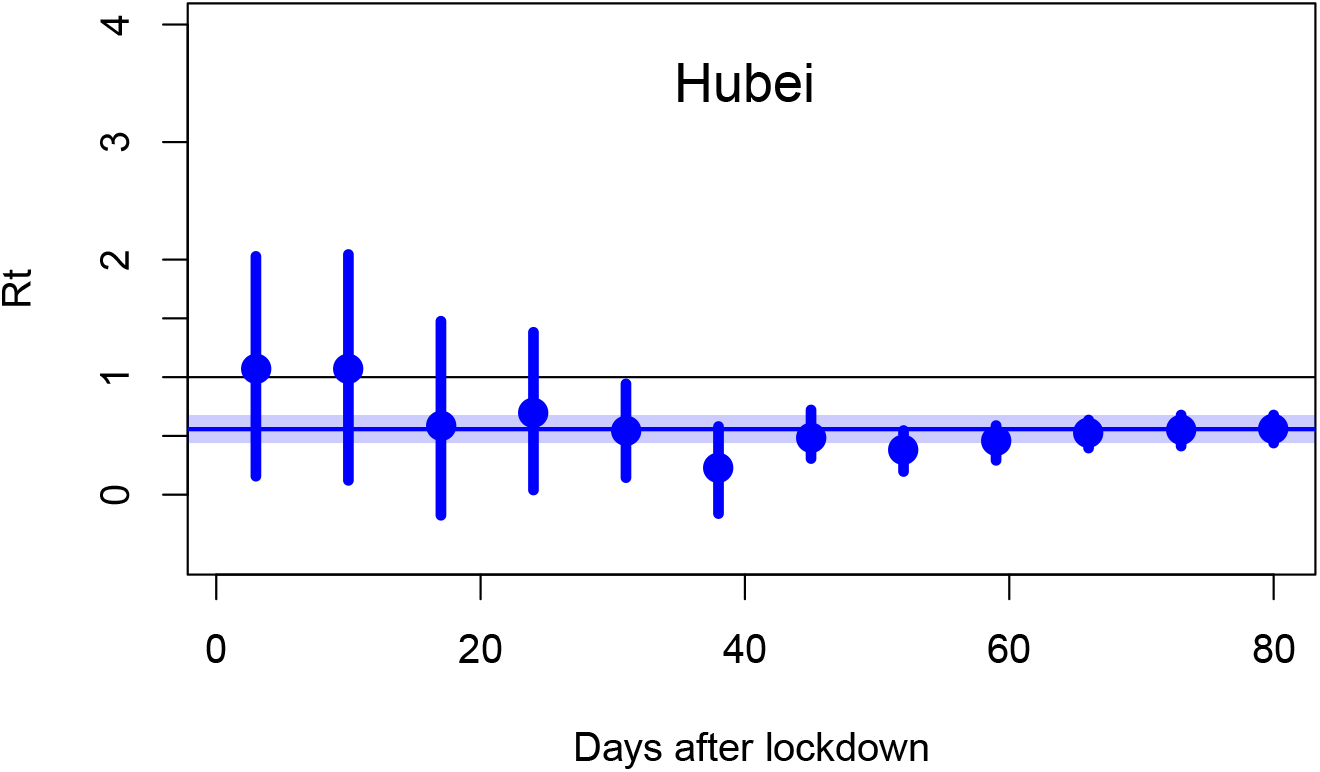
Estimates for Rt in Hubei Province. Median (dot) and 95% credible interval (line) estimates of *R_t_* at weekly intervals starting 3 days after lockdown data. Shaded region is 95% range of final estimate.

## UK and other European countries

We have been performing analyses for the UK on a daily basis (which have been published on Twitter: @jamesannan) and in Figure 5 we show a sample of these results generated at fortnightly intervals together with the most recent forecast at time of writing (16 May). The UK Government originally reported daily deaths occurring in hospital, but in late April the reporting protocol changed to include all deaths attributed to COVID-19 regardless of location. An implicit assumption of our analysis is that the reporting process is consistent, but fortunately at the time of the change in reporting standard the UK also generated a new historic time series of all attributed deaths and thus we present two sets of analyses for the UK, Figure 6A containing those based on the original hospital deaths up to late April, and Figure 6B containing a new set of analyses (including repeats of the early forecasts) based on all reported deaths. Despite the change in reporting standard, the results shown in Figures 6 show a high degree of consistency in estimated *R_t_* values during their period of overlap.

**Fig 5.**
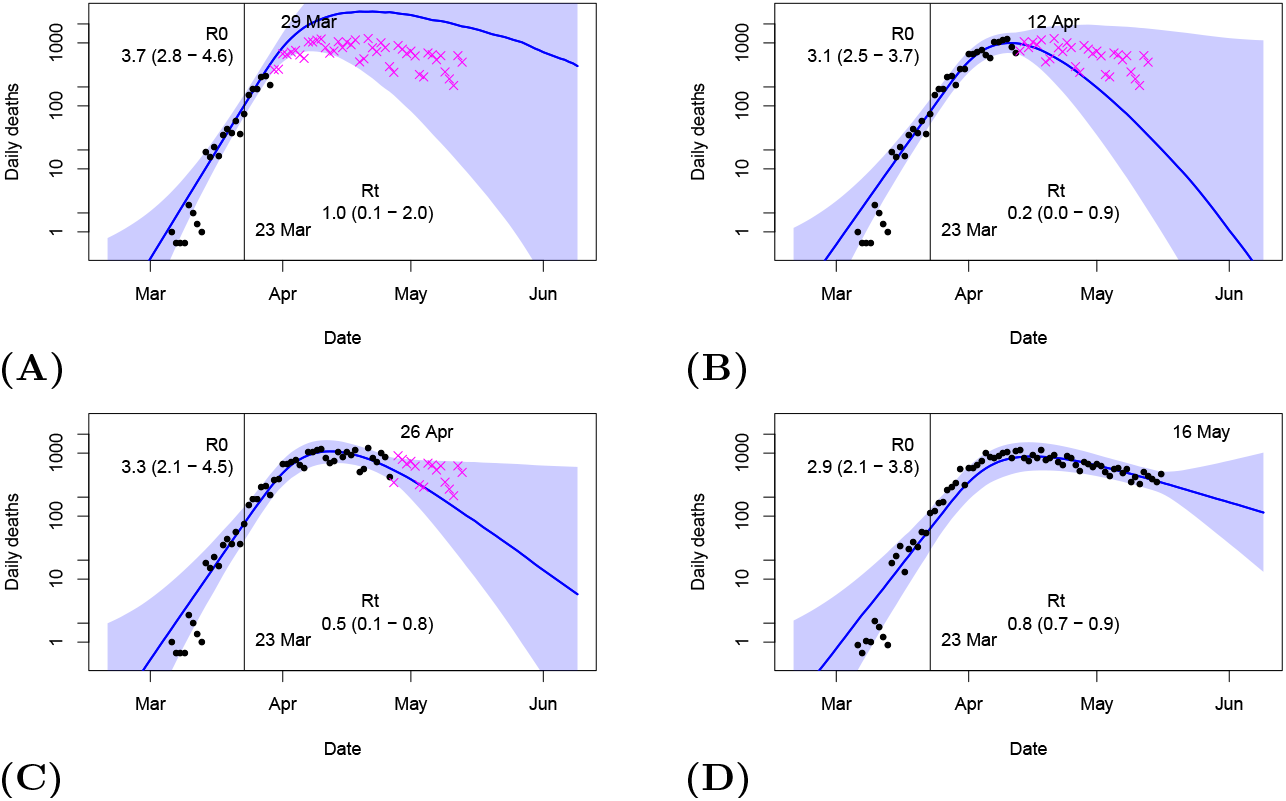
Hindcasts and forecasts for UK issued throughout epidemic. Median hindcast/forecast is shown as blue line, with blue plume as 95% credible interval. Date of lockdown shown as vertical line, with estimated *R*_0_ and *R_t_* values before and after this date provided as mean and 95% ranges. Black dots are daily deaths which were used for calibration of each forecast, with forecast date shown on plot. Magenta crosses are validation data observed subsequently to forecast date. Figures A–C based on hospital deaths as reported at the time, D based on all reported deaths

**Fig 6.**
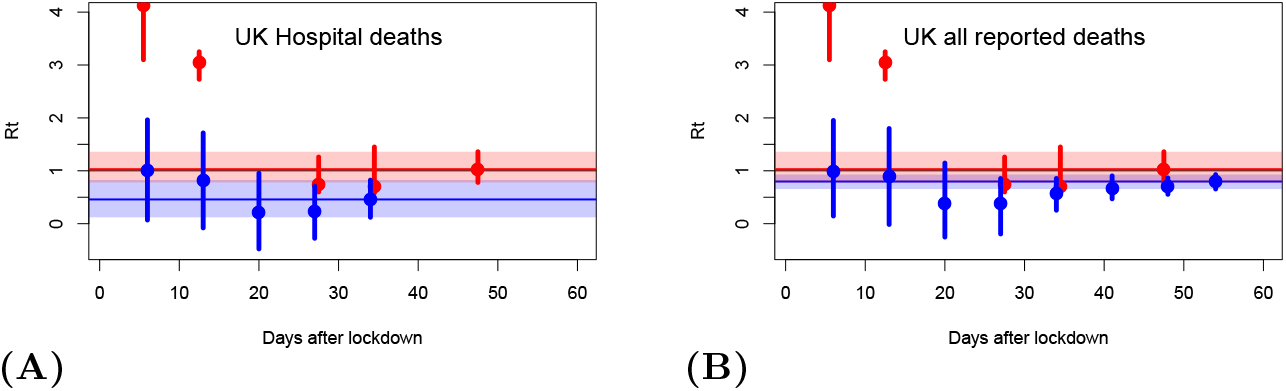
Estimates of *R_t_* for the UK. Analyses performed at weekly intervals after the date of lockdown (23 March). A hospital deaths and B all reported deaths. Blue dots and lines show median and 95% ranges from our analyses. Red dots and lines are equivalent results from MRC/IC analysis offset by half a day to avoid overlap. Pale blue and red bands show 95% ranges from the latest available analysis from each source.

Figure 7 shows equivalent results for three other European countries which experienced large outbreaks. These results are all broadly similar to the validation results shown in Figure 4 and show that the initial forecasts simply rely on the prior assumption for *R_t_* for around two weeks after the lockdown date, before converging smoothly to a value for *R_t_* of less than 1 which indicates successful suppression in each country.

**Fig 7.**
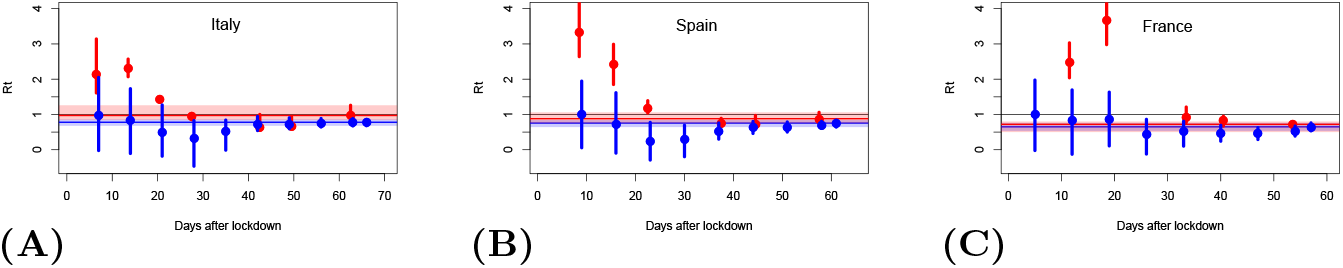
Estimates of *R_t_* for European countries. Analyses performed at weekly intervals after the date of lockdown (taken as 8, 13, and 17 March in A Italy, B Spain and C France respectively). Blue dots and lines show median and 95% ranges from our analyses. Red dots and lines are equivalent results from MRC/IC analysis. Pale blue and red bands show 95% ranges from the recent analysis plotted from each source.

In the UK, by April 11, which was less than 3 weeks from the lockdown date, we were able to correctly diagnose that the epidemic had been successfully sup-pressed with *R_t_* < 1 and that the peak in deaths had probably been passed [31], but expert opinion at that time incorrectly forecast rising death numbers for up to two more weeks [32].

Figures 6 and 7 include some other results of statistical analysis of death data. These analyses appear to have been made at weekly intervals and published on the web (https://mrc-ide.github.io/covid19-short-term-forecasts/index.html) but we were unable to obtain all of the historical forecasts. Their estimates of *R_t_* seem highly unstable and unreliable, with inconsistent and unrealistic confidence intervals in many cases. Their latest estimates (10 May 2020) still assign around 50% probability to values for *R_t_* greater than 1 for the UK and Spain, despite the steadily decreasing death numbers over the long term which according to our simulations can only be explained by *R_t_* < 1. These results seem surprising in the context of the long-term gradual decline in deaths that has been seen in all of these countries over the past several weeks.

The Institute for Health Metrics and Evaluation (IHME) also generated some predictions for the UK, which initially appear to have been based on fitting a Gaussian shape to historic data and extrapolated into the future [33]. Their first forecast for the UK did not validate. It was made on 12 April and predicted that the last deaths would occur by the 19th May. Their forecast on the 22nd April predicted that the epidemic would end at the start of June, which also seems unlikely. Subsequent to this, they changed their approach and are now making more plausible predictions based on an improved curve-fitting approach, with their most recent forecast being for a range of 40-50k deaths in total (for this wave) with the epidemic persisting to mid-July. Some other results based on statistical models [10] appear more reliable but have not been archived for comparison. We are unaware of any forecasts made by recognised research groups using calibrated dynamical models, and expect that such an approach using more sophisticated models would have outperformed our simple analysis. Therefore, we hope that our results will provoke further work in this area.

## Conclusion

We have shown that calibration of dynamical models is a powerful tool to help give credible policy-relevant support for the decision making process, and that this could easily be achieved in a fast-moving quasi-operational scenario such as the development of a new outbreak. We have presented a simple approach for calibrating a low complexity SEIR epidemic model using a Markov Chain Monte Carlo algorithm to perform Bayesian parameter estimation.

Calibrating the model to data available up to the 14th March obtains the rapid growth rate of 3 day doubling which continued up to the imposition of policy interventions. This approach generates greatly improved forecasts of the early growth of the epidemic, compared to model simulations which were presented by the Scientific Pandemic Influenza Group on Modelling (SPI-M) to the Scientific Advisory Group for Emergencies (SAGE) group at that time.

We also show that this method can detect the effectiveness of interventions to suppress the COVID-19 outbreak. Our approach detected that suppression (*R* < 1) had been successfully achieved in the UK and also several other European countries, earlier and more accurately than has been demonstrated with conventional statistical methods. Using this method, we were able to diagnose the peak of the daily deaths on the 11th April, two days after it occurred, and our current estimate for the reproduction number in the UK is 0.8 with a 95% credible range of 0.7–0.9.

## Data Availability

The datasets and code used for the current study are available from the corresponding author on reasonable request. Example code used for the analysis is available on GitHub.

https://github.com/epimath/cm-dag

## Code availability

Code for the generation of these analyses will be made available on Github: https://github.com/jdannan/COVID-19-operational-forecast. Daily nowcast/forecasts for the UK are published on Twitter: https://twitter.com/jamesannan.

## Acknowledgments

We appreciate numerous comments on our blog and also via Twitter which have helped greatly in improving the presentation of this work.

